## Supplement 1 for "Do people with disabilities experience disparities in cancer care? A systematic review"

#### - Search Strategy

**EMBASE AND MEDLINE**

((disabilit* or disable* or handicap* or function* limitation* or function* diversit* or dependen* or special need* or rare diseas* or capacit*) adj6 (person* or people or individ* or patient* or subject* or adult* or elderly)).ti,ab.

(Physical* adj5 (impair* or deficienc* or disable* or disabili* or handicap* or incapacit*)).ti,ab.

(Cerebral pals* or Spina bifida or Muscular dystroph* or Osteogenesis imperfecta or Musculoskeletal abnormalit* or Musculo-skeletal abnormalit* or Muscular abnormalit* or Skeletal abnormalit* or Limb abnormalit* or Amputation* or Clubfoot or Poliomyeliti* or Paraplegi* or Paralys* or Paralyz* or Hemiplegi* or wheelchair user* or wheel chair user*).ti,ab.

exp wheelchair user/

((Hearing or Acoustic or Ear or Ears) adj5 (loss* or impair* or deficienc* or disable* or disabili* or handicap*)).ti,ab.

((Visual* or Vision or Eye or eyes) adj5 (loss* or impair* or deficienc* or disable* or disabili* or handicap*)).ti,ab.

(Deaf* or Blind*).ti,ab.

exp Hearing impairment/

exp vision disorders/

(Schizophreni* or Psychos#s or Psychotic Disorder* or Schizoaffective Disorder* or Schizophreniform Disorder* or Dementia* or Alzheimer* or Bipolar Disorder* or personality disorder*).ti,ab.

exp "schizophrenia and disorders with psychotic features"/

exp Dementia/ or exp Alzheimer disease/

((Intellectual* or Mental* or Psychological* or Developmental or cognitive) adj5 (impair* or retard* or deficienc* or disable* or disabili* or handicap* or ill* or dysfunction* or deficit* or incapacit*)).ti,ab.

exp Mentally Disabled Persons/

((communication or language or speech or learning) adj5 (disorder* or disabilit* or impair* or deficit* or deficienc*)).ti,ab.

((genetic or hereditary or inherited or congenital) adj3 (disease* or ill* or syndrome or defect* or disorder* or condition* or malformation or anomal* or abnormalit*)).ti,ab.

exp *malignant neoplasm/co, dm, rt, rh, si, su, th [Complication, Disease Management, Radiotherapy, Rehabilitation, Side Effect, Surgery, Therapy]

(((disabilit* or disable* or handicap* or function* limitation* or function* diversit* or dependen* or special need* or rare diseas* or capacit*) adj6 (person* or people or individ* or patient* or subject* or adult* or elderly)) or (Physical* adj5 (impair* or deficienc* or disable* or disabili* or handicap* or incapacit*)) or (Cerebral pals* or Spina bifida or Muscular dystroph* or Osteogenesis imperfecta or Musculoskeletal abnormalit* or Musculo-skeletal abnormalit* or Muscular abnormalit* or Skeletal abnormalit* or Limb abnormalit* or Amputation* or Clubfoot or Poliomyeliti* or Paraplegi* or Paralys* or Paralyz* or Hemiplegi* or wheelchair user* or wheel chair user*) or wheelchair user or ((Hearing or Acoustic or Ear or Ears) adj5 (loss* or impair* or deficienc* or disable* or disabili* or handicap*)) or ((Visual* or Vision or Eye or eyes) adj5 (loss* or impair* or deficienc* or disable* or disabili* or handicap*)) or (Deaf* or Blind*) or Hearing impairment or vision disorders or (Schizophreni* or Psychos#s or Psychotic Disorder* or Schizoaffective Disorder* or Schizophreniform Disorder* or Dementia* or Alzheimer* or Bipolar Disorder* or personality disorder*) or "schizophrenia and disorders with psychotic features" or (Dementia or Alzheimer disease) or ((Intellectual* or Mental* or Psychological* or Developmental or cognitive) adj5 (impair* or retard* or deficienc* or disable* or disabili* or handicap* or ill* or dysfunction* or deficit* or incapacit*)) or Mentally Disabled Persons or ((communication or language or speech or learning) adj5 (disorder* or disabilit* or impair* or deficit* or deficienc*)) or ((genetic or hereditary or inherited or congenital) adj3 (disease* or ill* or syndrome or defect* or disorder* or condition* or malformation or anomal* or abnormalit*))).ti.

17 and 18

limit 19 to (human and english language and yr="2000 -Current" and (article or article in press) and journal and (adult <18 to 64 years> or aged <65+ years>))

limit 20 to ("young adult (19 to 24 years)" or "adult (19 to 44 years)" or "young adult and adult (19-24 and 19-44)" or "middle age (45 to 64 years)" or "middle aged (45 plus years)" or "all aged (65 and over)" or "aged (80 and over)") limit 21 to english language

limit 22 to human

limit 23 to (adult <18 to 64 years> or aged <65+ years>)

limit 24 to humans

limit 25 to (article or article in press or clinical study or clinical trial, all or clinical trial or controlled clinical trial or journal article or meta analysis or multicenter study or randomized controlled trial or "systematic review")

limit 26 to yr="2000 -Current"

limit 27 to journal

**COCHRANE**

ID Search Hits

#1 ((disabilit*:ti,ab OR disable*:ti,ab OR handicap*:ti,ab OR (function* NEXT limitation*):ti,ab OR (function* NEXT diversit*):ti,ab OR dependen*:ti,ab OR ("special" NEXT need*):ti,ab OR ("rare" NEXT diseas*):ti,ab OR capacit*:ti,ab) NEAR/6 (person*:ti,ab OR people:ti,ab OR individ*:ti,ab OR patient*:ti,ab OR subject*:ti,ab OR adult*:ti,ab OR elderly:ti,ab)) 30287

#2 (Physical*:ti,ab NEAR/5 (impair*:ti,ab OR deficienc*:ti,ab OR disable*:ti,ab OR disabili*:ti,ab OR handicap*:ti,ab OR incapacit*:ti,ab)) 3946

#3 (("Cerebral" NEXT pals*):ti,ab OR "Spina bifida":ti,ab OR ("Muscular" NEXT dystroph*):ti,ab OR "Osteogenesis imperfecta":ti,ab OR ("Musculoskeletal" NEXT abnormalit*):ti,ab OR ("Musculo-skeletal" NEXT abnormalit*):ti,ab OR ("Muscular" NEXT abnormalit*):ti,ab OR ("Skeletal" NEXT abnormalit*):ti,ab OR ("Limb" NEXT abnormalit*):ti,ab OR Amputation*:ti,ab OR Clubfoot:ti,ab OR Poliomyeliti*:ti,ab OR Paraplegi*:ti,ab OR Paralys*:ti,ab OR Paralyz*:ti,ab OR Hemiplegi*:ti,ab OR ("wheelchair" NEXT user*):ti,ab OR ("wheel chair" NEXT user*):ti,ab) 14521

#4 MeSH descriptor: [Wheelchairs] explode all trees 219

#5 ((Hearing:ti,ab OR Acoustic:ti,ab OR Ear:ti,ab OR Ears:ti,ab) NEAR/5 (loss*:ti,ab OR impair*:ti,ab OR deficienc*:ti,ab OR disable*:ti,ab OR disabili*:ti,ab OR handicap*:ti,ab)) 3453

#6 ((Visual*:ti,ab OR Vision:ti,ab OR Eye:ti,ab OR eyes:ti,ab) NEAR/5 (loss*:ti,ab OR impair*:ti,ab OR deficienc*:ti,ab OR disable*:ti,ab OR disabili*:ti,ab OR handicap*:ti,ab)) 6439

#7 (Deaf*:ti,ab OR Blind*:ti,ab) 351655

#8 MeSH descriptor: [Hearing Loss] explode all trees 1342

#9 MeSH descriptor: [Vision Disorders] explode all trees 1617

#10 (Schizophreni*:ti,ab OR Psychos?s:ti,ab OR ("Psychotic" NEXT Disorder*):ti,ab OR ("Schizoaffective" NEXT Disorder*):ti,ab OR ("Schizophreniform" NEXT Disorder*):ti,ab OR Dementia*:ti,ab OR Alzheimer*:ti,ab OR ("Bipolar" NEXT Disorder*):ti,ab OR ("personality" NEXT disorder*):ti,ab)45814

#11 MeSH descriptor: [Schizophrenia Spectrum and Other Psychotic Disorders] explode all trees 9836

#12 MeSH descriptor: [Dementia] explode all trees 6703

#13 MeSH descriptor: [Alzheimer Disease] explode all trees 3767

#14 ((Intellectual*:ti,ab OR Mental*:ti,ab OR Psychological*:ti,ab OR Developmental:ti,ab OR cognitive:ti,ab) NEAR/5 (impair*:ti,ab OR retard*:ti,ab OR deficienc*:ti,ab OR disable*:ti,ab OR disabili*:ti,ab OR handicap*:ti,ab OR ill*:ti,ab OR dysfunction*:ti,ab OR deficit*:ti,ab OR incapacit*:ti,ab)) 27248

#15 MeSH descriptor: [Persons with Mental Disabilities] explode all trees 62

#16 ((communication:ti,ab OR language:ti,ab OR speech:ti,ab OR learning:ti,ab) NEAR/5 (disorder*:ti,ab OR disabilit*:ti,ab OR impair*:ti,ab OR deficit*:ti,ab OR deficienc*:ti,ab)) 3421

#17 ((genetic:ti,ab OR hereditary:ti,ab OR inherited:ti,ab OR congenital:ti,ab) NEAR/3 (disease*:ti,ab OR ill*:ti,ab OR syndrome:ti,ab OR defect*:ti,ab OR disorder*:ti,ab OR condition*:ti,ab OR malformation:ti,ab OR anomal*:ti,ab OR abnormalit*:ti,ab)) 6887

#18 MeSH descriptor: [Neoplasms] explode all trees 88868

#19 #1 OR #2 OR #3 OR #4 OR #5 OR #6 OR #7 OR #8 OR #9 OR #10 OR #11 OR #12 OR #13 OR #14 OR #15 OR #16 OR #17 447483

#20 #18 AND 19 with Cochrane Library publication date Between Jan 2000 and Dec 2022, in Cochrane Reviews, Trials 614

**WEB OF SCIENCE**

https://www.webofscience.com/wos/woscc/summary/2b2baa98-ffbf-4d8b-a325-d948689254ea-43442f08/relevance/1

**CINAHL**

(((TI disabilit* OR AB disabilit*) OR (TI disable* OR AB disable*) OR (TI handicap* OR AB handicap*) OR (TI "function* limitation*" OR AB "function* limitation*") OR (TI "function* diversit*" OR AB "function* diversit*") OR (TI dependen* OR AB dependen*) OR (TI "special need*" OR AB "special need*") OR (TI "rare diseas*" OR AB "rare diseas*") OR (TI capacit* OR AB capacit*)) N6 ((TI person* OR AB person*) OR (TI people OR AB people) OR (TI individ* OR AB individ*) OR (TI patient* OR AB patient*) OR (TI subject* OR AB subject*) OR (TI adult* OR AB adult*) OR (TI elderly OR AB elderly)))

((TI Physical* OR AB Physical*) N5 ((TI impair* OR AB impair*) OR (TI deficienc* OR AB deficienc*) OR (TI disable* OR AB disable*) OR (TI disabili* OR AB disabili*) OR (TI handicap* OR AB handicap*) OR (TI incapacit* OR AB incapacit*)))

((TI "Cerebral pals*" OR AB "Cerebral pals*") OR (TI "Spina bifida" OR AB "Spina bifida") OR (TI "Muscular dystroph*" OR AB "Muscular dystroph*") OR (TI "Osteogenesis imperfecta" OR AB "Osteogenesis imperfecta") OR (TI "Musculoskeletal abnormalit*" OR AB "Musculoskeletal abnormalit*") OR (TI "Musculo-skeletal abnormalit*" OR AB "Musculo-skeletal abnormalit*") OR (TI "Muscular abnormalit*" OR AB "Muscular abnormalit*") OR (TI "Skeletal abnormalit*" OR AB "Skeletal abnormalit*") OR (TI "Limb abnormalit*" OR AB "Limb abnormalit*") OR (TI Amputation* OR AB Amputation*) OR (TI Clubfoot OR AB Clubfoot) OR (TI Poliomyeliti* OR AB Poliomyeliti*) OR (TI Paraplegi* OR AB Paraplegi*) OR (TI Paralys* OR AB Paralys*) OR (TI Paralyz* OR AB Paralyz*) OR (TI Hemiplegi* OR AB Hemiplegi*) OR (TI "wheelchair user*" OR AB "wheelchair user*") OR (TI "wheel chair user*" OR AB "wheel chair user*"))

(MH "wheelchair user"+)

(((TI Hearing OR AB Hearing) OR (TI Acoustic OR AB Acoustic) OR (TI Ear OR AB Ear) OR (TI Ears OR AB Ears)) N5 ((TI loss* OR AB loss*) OR (TI impair* OR AB impair*) OR (TI deficienc* OR AB deficienc*) OR (TI disable* OR AB disable*) OR (TI disabili* OR AB disabili*) OR (TI handicap* OR AB handicap*)))

(((TI Visual* OR AB Visual*) OR (TI Vision OR AB Vision) OR (TI Eye OR AB Eye) OR (TI eyes OR AB eyes)) N5 ((TI loss* OR AB loss*) OR (TI impair* OR AB impair*) OR (TI deficienc* OR AB deficienc*) OR (TI disable* OR AB disable*) OR (TI disabili* OR AB disabili*) OR (TI handicap* OR AB handicap*)))

((TI Deaf* OR AB Deaf*) OR (TI Blind* OR AB Blind*))

(MH "Hearing impairment"+)

(MH "vision disorders"+)

((TI Schizophreni* OR AB Schizophreni*) OR (TI Psychos?s OR AB Psychos?s) OR (TI "Psychotic Disorder*" OR AB "Psychotic Disorder*") OR (TI "Schizoaffective Disorder*" OR AB "Schizoaffective Disorder*") OR (TI "Schizophreniform Disorder*" OR AB "Schizophreniform Disorder*") OR (TI Dementia* OR AB Dementia*) OR (TI Alzheimer* OR AB Alzheimer*) OR (TI "Bipolar Disorder*" OR AB "Bipolar Disorder*") OR (TI "personality disorder*" OR AB "personality disorder*"))

(MH "schizophrenia and disorders with psychotic features"+)

(MH Dementia+) OR (MH "Alzheimer disease"+)

(((TI Intellectual* OR AB Intellectual*) OR (TI Mental* OR AB Mental*) OR (TI Psychological* OR AB Psychological*) OR (TI Developmental OR AB Developmental) OR (TI cognitive OR AB cognitive)) N5 ((TI impair* OR AB impair*) OR (TI retard* OR AB retard*) OR (TI deficienc* OR AB deficienc*) OR (TI disable* OR AB disable*) OR (TI disabili* OR AB disabili*) OR (TI handicap* OR AB handicap*) OR (TI ill* OR AB ill*) OR (TI dysfunction* OR AB dysfunction*) OR (TI deficit* OR AB deficit*) OR (TI incapacit* OR AB incapacit*)))

(MH "Mentally Disabled Persons"+)

(((TI communication OR AB communication) OR (TI language OR AB language) OR (TI speech OR AB speech) OR (TI learning OR AB learning)) N5 ((TI disorder* OR AB disorder*) OR (TI disabilit* OR AB disabilit*) OR (TI impair* OR AB impair*) OR (TI deficit* OR AB deficit*) OR (TI deficienc* OR AB deficienc*)))

(((TI genetic OR AB genetic) OR (TI hereditary OR AB hereditary) OR (TI inherited OR AB inherited) OR (TI congenital OR AB congenital)) N3 ((TI disease* OR AB disease*) OR (TI ill* OR AB ill*) OR (TI syndrome OR AB syndrome) OR (TI defect* OR AB defect*) OR (TI disorder* OR AB disorder*) OR (TI condition* OR AB condition*) OR (TI malformation OR AB malformation) OR (TI anomal* OR AB anomal*) OR (TI abnormalit* OR AB abnormalit*)))

(MM "malignant neoplasm"/co,+) "dm, rt, rh, si, su, th [Complication, Disease Management, Radiotherapy, Rehabilitation, Side Effect, Surgery, Therapy]"

(((TI disabilit* OR TI disable* OR TI handicap* OR TI "function* limitation*" OR TI "function* diversit*" OR TI dependen* OR TI "special need*" OR TI "rare diseas*" OR TI capacit*) N6 (TI person* OR TI people OR TI individ* OR TI patient* OR TI subject* OR TI adult* OR TI elderly)) OR (TI Physical* N5 (TI impair* OR TI deficienc* OR TI disable* OR TI disabili* OR TI handicap* OR TI incapacit*)) OR (TI "Cerebral pals*" OR TI "Spina bifida" OR TI "Muscular dystroph*" OR TI "Osteogenesis imperfecta" OR TI "Musculoskeletal abnormalit*" OR TI "Musculo-skeletal abnormalit*" OR TI "Muscular abnormalit*" OR TI "Skeletal abnormalit*" OR TI "Limb abnormalit*" OR TI Amputation* OR TI Clubfoot OR TI Poliomyeliti* OR TI Paraplegi* OR TI Paralys* OR TI Paralyz* OR TI Hemiplegi* OR TI "wheelchair user*" OR TI "wheel chair user*") OR TI "wheelchair user" OR ((TI Hearing OR TI Acoustic OR TI Ear OR TI Ears) N5 (TI loss* OR TI impair* OR TI deficienc* OR TI disable* OR TI disabili* OR TI handicap*)) OR ((TI Visual* OR TI Vision OR TI Eye OR TI eyes) N5 (TI loss* OR TI impair* OR TI deficienc* OR TI disable* OR TI disabili* OR TI handicap*)) OR (TI Deaf* OR TI Blind*) OR TI "Hearing impairment" OR TI "vision disorders" OR (TI Schizophreni* OR TI Psychos?s OR TI "Psychotic Disorder*" OR TI "Schizoaffective Disorder*" OR TI "Schizophreniform Disorder*" OR TI Dementia* OR TI Alzheimer* OR TI "Bipolar Disorder*" OR TI "personality disorder*") OR TI "schizophrenia and disorders with psychotic features" OR (TI Dementia OR TI "Alzheimer disease") OR ((TI Intellectual* OR TI Mental* OR TI Psychological* OR TI Developmental OR TI cognitive) N5 (TI impair* OR TI retard* OR TI deficienc* OR TI disable* OR TI disabili* OR TI handicap* OR TI ill* OR TI dysfunction* OR TI deficit* OR TI incapacit*)) OR TI "Mentally Disabled Persons" OR ((TI communication OR TI language OR TI speech OR TI learning) N5 (TI disorder* OR TI disabilit* OR TI impair* OR TI deficit* OR TI deficienc*)) OR ((TI genetic OR TI hereditary OR TI inherited OR TI congenital) N3 (TI disease* OR TI ill* OR TI syndrome OR TI defect* OR TI disorder* OR TI condition* OR TI malformation OR TI anomal* OR TI abnormalit*)))

S17 AND S18

"limit 19 to" (human AND "english language" AND "yr="2000 -Current"" AND (article OR "article in press" ) AND journal AND (adult UNKNOWN-TEMPLATE:18 to 64 years OR aged UNKNOWN-TEMPLATE:65+ years))

"limit 20 to" ("young adult (19 to 24 years)" OR "adult (19 to 44 years)" OR "young adult and adult (19-24 and 19-44)" OR "middle age (45 to 64 years)" OR "middle aged (45 plus years)" OR "all aged (65 and over)" OR "aged (80 and over)" )

"limit 21 to english language"

"limit 22 to human"

"limit 23 to" (adult UNKNOWN-TEMPLATE:18 to 64 years OR aged UNKNOWN-TEMPLATE:65+ years)

"limit 24 to humans"

"limit 25 to" (article OR "article in press" OR "clinical study" OR "clinical trial, all" OR "clinical trial" OR "controlled clinical trial" OR "journal article" OR "meta analysis" OR "multicenter study" OR "randomized controlled trial" OR "systematic review" ) "limit 26 to yr="2000 -Current""

"limit 27 to journal"

---

#### Determining risk of bias

For all study designs:

- Study design, sampling method was appropriate to the study question
- Adequate sample size
- Response rate was reported and acceptable (>70%)
- Disability/impairment measure was clearly defined and reliable
- Measure of outcome was clearly defined and reliable
- Potential confounders were taken into account in analysis

- Confidence intervals were presented

Additional criteria for case control studies:

- Cases and controls were comparable
- Cases and controls were clearly defined

Additional criteria for cohort studies:

- Groups being studied were comparable at baseline
- Losses to follow up were presented and acceptable

### Table 1. Details of study characteristics

| AUTHOR | YEAR | COUNTRY | STUDY DESIGN | CANCER TYPE | N WITH DISABILITIES | N WITHOUT | DISABILITY TYPE | PRIMARY OUTCOME | SECONDARY OUTCOME | RISK OF BIAS |
|---|---|---|---|---|---|---|---|---|---|---|
| Afshar(53) | 2020 | UK | Retrospective cohort | Testicular | 331 | 25675 | Intellectual (Learning disability) | 10-yr survival rate | 5-yr survival rate | Medium |
| Chang(51) | 2013 | Taiwan | Retrospective cohort | Oral cancer | 206 | 16481 | Psychosocial (Mental illness) | Access to state-of-the-art treatment | Mortality | Medium |
| Cuypers(56) | 2022 | Netherlands | Retrospective cohort | Any cancer | 187149 | 12677768 | Intellectual | Cancer-specific mortality | n/a | Medium |
| Cuypers(70) | 2020 | Netherlands | Retrospective cohort | Any cancer as inpatient | 65183 | 129497 | Intellectual | Insurance claims for cancer hospital care | n/a | Medium |

| | | | | | | | | | | |
|---|---|---|---|---|---|---|---|---|---|---|
| Fond (40) | 2021 | France | Retrospective cohort | Breast | 1742 | 36870 | Psychosocial (Severe psychiatric disease) | End-of-life treatment access | Overall survival | Medium |
| Fried(41) | 2019 | USA | Retrospective cohort | Nonmetastatic prostate | 523 | 49462 | Psychosocial (Severe mental illness) | Cancer-specific 5-yr mortality | Access to state-of-the-art treatment | Low |
| Gross(63) | 2020 | Germany | Patient survey | Breast | 568 | 4058 | Any disability | Screening results | Invasiveness of treatment | High |
| Gupta(71) | 2004 | USA | Retrospective cohort | Colon | 1184 | 16323 | Cognitive (Dementia) | Stage at diagnosis | Access to state-of-the-art treatment | Medium |
| Iezzoni(62) | 2008 | USA | Retrospective cohort | Non-small cell lung cancer | 1016 | 8425 | Any disability | Cancer-specific mortality | Access to state-of-the-art treatment | Low |
| Iglay(43) | 2017 | USA | Retrospective cohort | Breast | 3961 | 12675 | Psychosocial (Mental illness) | Treatment delay | Diagnosis delay | Medium |
| Ishikawa(42) | 2016 | Japan | Retrospective cohort | Gastric or colorectal | 2495 | 6521 | Psychosocial (Schizophrenia) | Overall mortality | Stage at diagnosis and | Low |

| | | | | | | | | | | |
|---|---|---|---|---|---|---|---|---|---|---|
| | | | | | | | | | access to state-of-the-art treatment | |
| Kaneshiro(44) | 2021 | Japan | Retrospective cohort | Breast | 55 | 610 | Psychosocial (Schizophrenia) | Invasiveness of treatment | Access to state-of-the-art treatment | High |
| Kashyap(45) | 2021 | USA | Retrospective cohort | Gastro-intestinal | 54661 | 105706 | Psychosocial (Mental illness) | End of life Emergency Department use | Impact of outpatient mental health treatment in mental illness | Low |
| Kim(22) | 2020 | South Korea | Retrospective cohort | Stomach | 16849 | 36852 | Any disability | Mortality | Survival after surgery | Low |
| Kwon(19) | 2020 | South Korea | Retrospective cohort | Multiple Myeloma (MM) | 809 | 3281 | Any disability | Overall survival | Access to state-of-the-art treatment | Low |
| Kwon(68) | 2020 | South Korea | Retrospective cohort | Acute Myeloid Leukemia (AML) | 566 | 2227 | Any disability | Overall survival | Access to state-of-the-art treatment | Low |

| | | | | | | | | | | |
|---|---|---|---|---|---|---|---|---|---|---|
| Lawrence(67) | 2020 | USA | Retrospective cohort | Breast | 3203 | 7201 | Psychosocial (Severe mental illness) | All-cause and cancer-specific mortality | 10-year overall survival | Medium |
| Libert(61) | 2016 | Belgium | Prospective cohort | Breast, prostate, colorectal | 164 | 193 | Cognitive | Overall mortality | n/a | Medium |
| Mahabaleshwarkar(47) | 2015 | USA | Retrospective cohort | Breast | 806 | 1306 | Psychosocial (Mental illness) | Access to state-of-the-art treatment | Healthcare utilization | Medium |
| Martin(54) | 2020 | UK | Prospective cohort | Operable breast | 478 | 2938 | Cognitive | Overall mortality | Access to state-of-the-art treatment | Medium |
| Park(66) | 2011 | South Korea | Retrospective cohort | Any cancer | 4077 | 89681 | Any disability | Long-term all-cause mortality of 5-year cancer survivors | Short-term (<5 years) all-cause mortality | Low |

| | | | | | | | | | | |
|---|---|---|---|---|---|---|---|---|---|---|
| Robb(57) | 2009 | USA | Retrospective case-control | Any cancer | 86 | 172 | Cognitive | Survival | n/a | High |
| Sathianathen(69) | 2019 | USA | Retrospective cohort | Bladder | 5194 | 62008 | Psychosocial (Mental illness) | Access to state-of-the-art treatment | Cancer-specific mortality | Medium |
| Sato(64) | 2021 | Japan | Retrospective cohort | Any cancer | 2983 | 90562 | Any disability | Access to state-of-the-art treatment | n/a | High |
| Segerlantz(59) | 2019 | Sweden | Retrospective cohort | Any cancer | 555 | 877 | Intellectual | Pain control prescription | Prescription of other drugs | Low |
| Segerlantz(58) | 2020 | Sweden | Retrospective cohort | Any cancer | 775 | 2968 | Intellectual | Healthcare utilization | Quality of end-of-life care | Low |
| Shin(68) | 2018 | South Korea | Retrospective cohort | Lung | 13591 | 43809 | Any disability | Overall mortality | Access to state-of-the-art treatment | Low |
| Shin(65) | 2020 | South Korea | Retrospective cohort | Prostate | 7924 | 34188 | Any disability | Access to state-of-the-art treatment | Overall mortality and cancer-specific mortality | Low |

| | | | | | | | | | | |
|---|---|---|---|---|---|---|---|---|---|---|
| Shinden(49) | 2017 | Japan | Retrospective cohort | Breast | 46 | 727 | Psychosocial (Mental illness) | Access to state-of-the-art treatment | Overall survival | High |
| Tran(50) | 2009 | France | Prospective cohort | Any cancer | 3400 | sample of general population | Psychosocial (Schizophrenia) | All-cause and all-cancer-mortality | Mortality by cancer type | Medium |
| Viprey(46) | 2020 | France | Retrospective cohort | Terminal lung cancer | 633 | 66469 | Psychosocial (Schizophrenia) | Access to state-of-the-art treatment | Quality of end-of-life care | Medium |

LEGEND:

yr = year; CI = Confidence Interval; OR = Odds Ratio; SMR = Standardized Mortality Ratio; IR = Incidence Rate; IRR = Incidence Rate Ratio; aOR = adjusted Odds Ratio; HR = Hazard Ratio; PWD = People with disabilities; aRR = adjusted Risk Ratio; ED = Emergency Department; aHR = adjusted Hazard Ratio; SMI = Severe Mental Illness; aIRR = adjusted Incidence Rate Ratio; PET = Primary Endocrine Treatment; NMIBC = non-muscle invasive bladder cancer; MIBC = muscle invasive bladder cancer; COX = cyclooxygenase; RR = Relative Risk; ADT = Androgen Deprivation Therapy; ICU = Intensive Care Unit
